# Parity and cumulative incidence rates of breast cancer in the Norwegian Woman and Cancer Study (NOWAC)

**DOI:** 10.1101/2024.10.10.24315223

**Authors:** Sanda Krum-Hansen, Arne Bastian Wiik, Karina Standahl Olsen, Marko Lukic, Ruth H. Paulssen, Eiliv Lund

## Abstract

**Background:** The reduced risk of breast cancer (BC) following increasing parity has been known for decades. Most prospective studies have presented the relative risk as the percentage decrease for each child during follow-up. Since the risk reduction is up to ten percent for each child, the overall lifelong BC risk reduction could be under communicated. In this study we use cumulative incidence rates (CIR) to calculate and describe the lifelong risk of BC in relation to parity.

**Methods:** NOWAC is a prospective cohort study with 172,000 women recruited between 1991 and 2007 with follow-up through questionnaires and national registers of cancer and death. For the present analyses, we included 165 238 women with follow-up from 01.01.2000 until 31.12. 2018. We calculated CIR of BC by parity, stratified by other established BC risk factors (maternal age at first birth, breastfeeding, body mass index (BMI), smoking and alcohol consumption).

**Results:** After 17.3 years of average follow-up, 8120 women aged 35-84 years developed breast cancer. Age-specific incidence rates increased for each age group up to 60-64 years, decreased for the age group 75-79 years, and increased again among the oldest women aged 80-84. CIR for all participants up to 84 years was 11 700 per 100 000 person years (PY). In analyses stratified by parity, the CIR of BC for nullipara was 12 600 per 100 000 PY, for 1-2 children: 12 100, 3-4 children: 10 200, and 5-6 children: 8 700 per 100 000 PY. The parity-specific CIR of BC had the same pattern of decrease in analyses stratified for other BC risk factors.

**Conclusion:** Cumulative incidence rates showed a consistent decrease in BC risk for each additional child. The decrease was consistent in strata of other established BC risk factors.

## 1 Introduction

Breast cancer (BC) is the most common cancer among women, and the leading cause of cancer death in women worldwide (1). Globally, BC incidence has increased over the last three decades (2), and the same trend has been observed in European countries, including Norway (3).

A large number of risk factors for developing BC have been studied: reproductive history (age at first birth, parity and duration of breastfeeding), body mass index (BMI), physical activity, diet including fat intake, alcohol consumptions, smoking, air pollution and radiation (4). Reproductive history and length of exposure to endogenous hormones (age at menarche, age at menopause) have a strong influence on the risk of developing BC (5). Exposure to exogenous hormones, use of oral contraceptive and hormonal therapy, lead to a transient increased risk during use, and 10 years, respective 2 years after cessation (6, 7).

Numerous epidemiological studies of parity have provided evidence of higher risk of BC in nulliparous compared with parous women, and declining incidence of BC with increasing number of children (8, 9). In many large prospective cohort studies and meta-analyses, the protective effect of parity for development of postmenopausal BC is around 7-8 % for every full-term pregnancy (9–12). Prolonged breast feeding provides additional risk reduction (13, 14).

Most epidemiological studies have used relative risk (RR) as a measure for association between exposure (parity) and outcome (BC). RR shows the risk of outcome over the follow-up time but tells little about lifetime risk. To our knowledge, only two previous study used CIR to investigate lifetime risk of BC (15, 16) neither of which calculated the change in CIR of BC for each pregnancy.

The aim of this study was to describe the parity-related lifetime risk of BC in a large prospective cohort by using CIR, and to investigate the parity-specific CIR of BC in strata of others BC risk factors.

## 2 Material and methods

### 2.1 Study design and participants

In this study we used data from The Norwegian Women and Cancer Study (NOWAC) and from the national registers of cancer and death. The NOWAC study (17) is a population-based prospective cohort study in which a sample of women living in Norway was recruited between 1991 and 2007 by random sampling from the Norwegian Central Population Registry. A total of 172 748 women participated with at least one questionnaire. Disease status, death and emigration status were updated through linkage to the Cause of Death Registry at Statistics Norway and to the Cancer Registry of Norway, using the national personal identification number. The exit date was defined as either the date of cancer diagnosis, death, emigration, or end of follow-up on December 31^st^, 2018. The date of entry into the present study was January 1^st^, 2000, because in 2000, most large counties in Norway had started mammography screening for BC. Since the screening was anticipated to increase the incidence rates in women aged 50-65 years, use of information from before 2000 could be difficult to interpret. After excluding prevalent cases of breast cancer and women with 7 children or more (for data anonymization purposes), 165 238 Norwegian women were included in analyses.

BC cases were identified through the Norwegian Cancer Registry identified according to organ site code C50 in the International Classification of Diseases, Tenth Revision (ICD-10) (https://icd.who).

### 2.2 Questionnaire information

Information on parity and other covariates was collected from the NOWAC baseline questionnaire. Parity was analyzed both as discrete numbers in the range from zero to six, and as four parity groups 0, 1-2, 3-4 and 5-6 children, respectively.

Breastfeeding was self-reported in months of duration per child and calculated by summing the total duration of breastfeeding and dividing by number of children. In the analyses, women who breastfed ≥6 months per child were compared to those who breastfed less than 6 months per child. Maternal age at first birth was calculated by the year of birth of the first child.

BMI (kg/m^2^) was calculated from self-reported data on weight and height. The variable was dichotomized into BMI<25 and BMI≥25. The smoking status in NOWAC was defined as current, former, or never smokers. We combined current and former smokers into a single category of ever-smokers. Alcohol consumption was defined as ever-drinkers and teetotalers.

The age range for the analyses of parity was 40-84 years. However, in the analyses stratified according to other BC risk factors, this was reduced to 40-79 years due to few BC cases in women aged 80-84. In the analyses stratified by alcohol use, we also excluded age group 75-79 years for parity group 5-6 children due to zero cases in alcohol users’ strata.

An additional change was done in analyses stratified by maternal age at the first birth; by taking into account the age of menopause, the youngest age group was changed from 40-45 to 45-50 to capture complete fertility history. For analyses stratified by breastfeeding, the number of parity groups was changed from three groups (0, 1–2, 3–4, 5–6) to two groups (1-3 and 4-6), due to few cases in each parity group.

### 2.3 Statistical methods

Characteristics of the study population overall and by BC risk factors were described as means with standard deviation (SD) and absolute numbers with percentages.

The overall and parity-specific CIR was calculated for each age-group as the number of incident cases of BC divided by the number of person-years (per 100 000 PY), until death, emigration, or a BC diagnosis. CIRs for lifestyle exposures per age group were calculated for BMI, smoking status and alcohol consumption.

The same calculation was done for the parity-specific CIR of BC, stratified by other BC risk factors: maternal age at first birth (before or after 25 years of age), duration of breast feeding (more or less than 6 months), BMI (above or below 25), alcohol consumption (yes or no), smoking status (ever or never). For all analyses of parity-specific CIR stratified by other BC risk factors, parity was categorized into groups.

The statistical software used was R statistical environment and the Epi package for Epidemiological research (R core team, 2022).

## 3 Results

### 3.1 Baseline characteristics of study population

The mean age at enrollment for all the women in the population was 49.5 years. The mean number of children was 2.2 (SD 1.2) and the mean maternal age at first birth was 23.8 years (SD 4.2). The average follow-up time was 17.3 years. During the follow-up period, 8120 cases of breast cancer were diagnosed.

Table 1 shows the distribution of exposures and baseline characteristics of the total study population and according to parity groups. One third of the total study population had BMI ≥25, most pronounced among those with high parity. The prevalence of alcohol consumption and smoking showed no clear trend with parity. Age at first birth showed a very strong gradient – women with higher parity have their first child at a younger age.

**Table 1.** Characteristics of the Study Population by Parity and Breast Cancer Risk Factors in the Norwegian Women and Cancer Study.

| Characteristics of study population | Total | Parity |  |  |  | Missing, n (%) |
| --- | --- | --- | --- | --- | --- | --- |
|  |  | 0 | 1-2 children | 3-4 children | 5-6 children |  |
| Number of women, n (%) | 164 105 | 16 124 | 88 243 | 55 280 | 4458 | 0 |
| Age at enroll., mean (SD) | 49.4 (8.4) | 48.8 (8.9) | 48.7 (8.2) | 50.3 (8.4) | 55.2 (8.4) | 0 |
| Age at 1st birth, n (%) |  |  |  |  |  |  |
| <25 years | 89 945 (54.81%) | N/A | 46 643 (52.86%) | 39 604 (71.64%) | 3698 (82.95%) | 244 (0.15%) |
| ≥25 years | 57 792 (35.22%) | N/A | 41 420 (46.94%) | 15 621 (28.26%) | 751 (16.85%) |  |
| Avg. months of breastfeeding per child, n (%) |  |  |  |  |  |  |
| <6 months | 39 937 (27%) | N/A | 30 794 (34.9%) | 8 624 (15.6%) | 519 (11.6%) | 0 |
| ≥6 months | 108 044 (73.0%) | N/A | 57 449 (65.1%) | 46 656 (84.4%) | 3 939 (88.4%) |  |
| BMI, n (%) |  |  |  |  |  |  |
| ≤25 | 104 529 (63.70%) | 10 448 (64.80%) | 58 374 (66.15%) | 33 606 (60.79%) | 2101 (47.13%) | 4075 (2.48%) |
| >25 | 55 501 (33.82%) | 5676 (35.20%) | 29 869 (33.85%) | 21 674 (39.21%) | 2357 (52.87%) |  |
| Alcohol use, n (%) |  |  |  |  |  |  |
| Yes | 93 215 (56.8%) | 9374 (58.14%) | 54 721 (62.01%) | 27 851 (50.38%) | 1269 (28.47%) | 8839 (5.39%) |
| No | 62 051 (37.81%) | 5792 (35.2%) | 29 244 (33.14%) | 24 205 (43.79%) | 2810 (63.03%) |  |
| Smoking status, n (%) |  |  |  |  |  |  |
| Ever | 105 912 (64.54%) | 9016 (55.92%) | 59 825 (67.8%) | 33 906 (61.34%) | 2350 (52.71%) | 2208 (1.35%) |
| Never | 55 985 (34.12%) | 6016 (37.31%) | 27 340 (30.98%) | 20 613 (37.29%) | 2016 (45.22%) |  |

Abbreviations: BMI, body mass index. SD, standard deviation.

### 3.2 Parity-specific CIR of BC

Age-specific incidence rates for BC in the NOWAC study increased from age 35-39 up to 60-64 years, decreased until 75-79 years, with another increase in the oldest age group (Figure 1a). Overall CIR for breast cancer in NOWAC was comparable to national levels (Figure 1b).

**Figure 1a.**
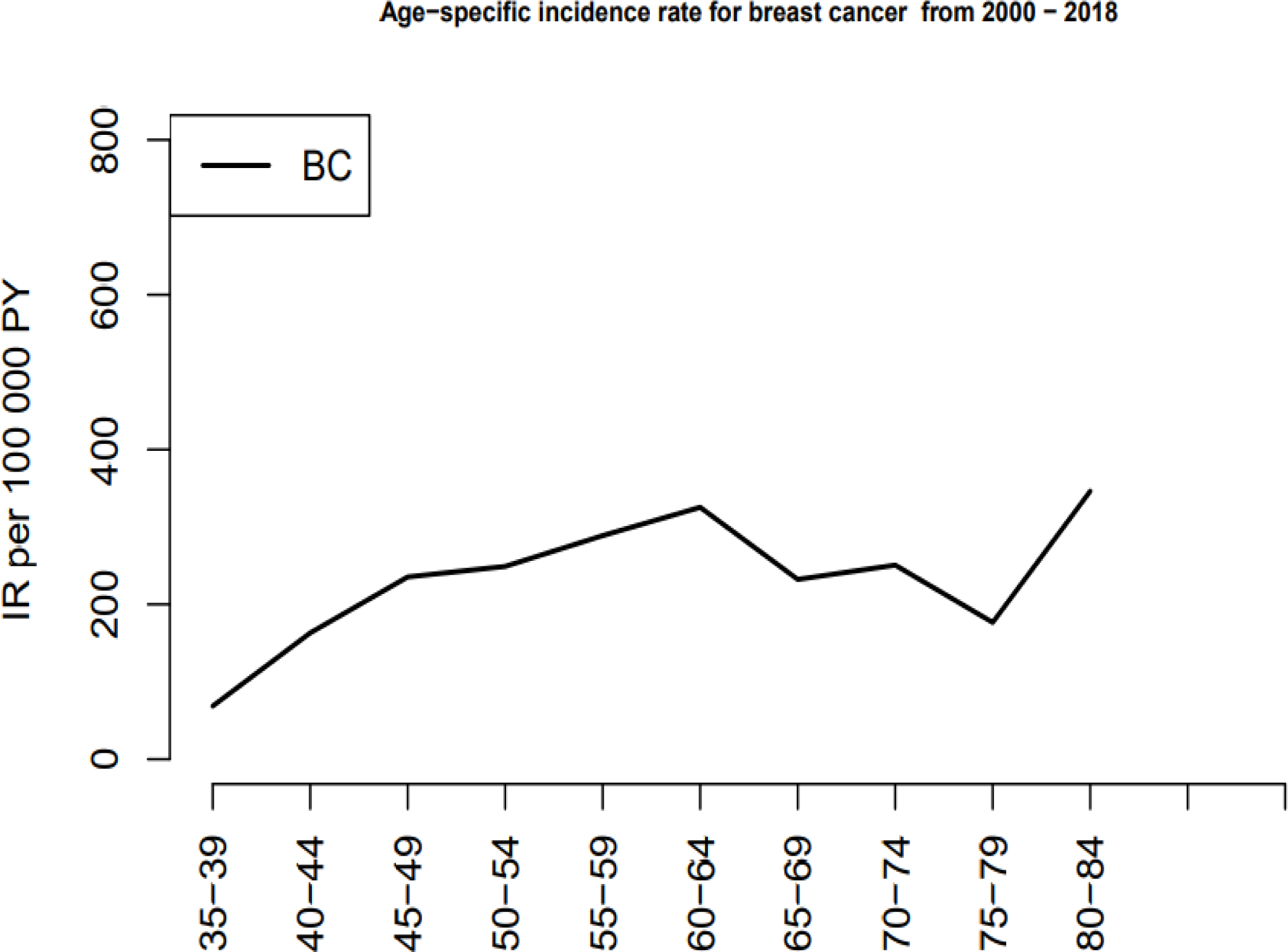
Age-specific incidence rates in NOWAC 2000-2018.

**Figure 1b.**
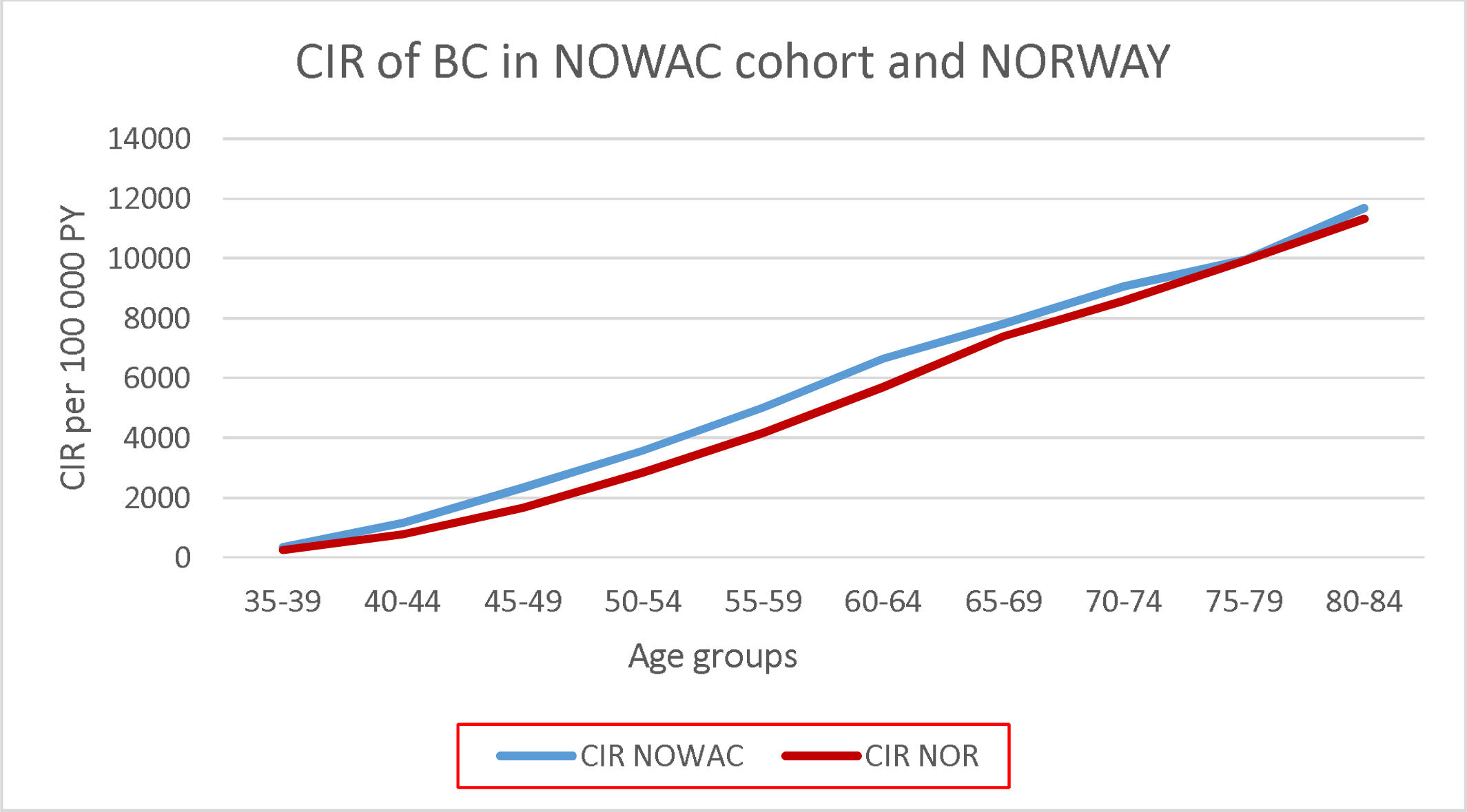
Cumulative incidence rate of breast cancer in the NOWAC cohort compared to national figures for the same years based on the Cancer Registry of Norway.

For nullipara, the CIR for all age groups were 12 600 per 100 000 PY. CIR for 1-2 children was 12 100, for 3-4 children CIR was 10 200, and for 5-6 children CIR was 8700 per 100 000 PY. (Table 2). Plots (Figure 2) show a negative association between parity and the age-specific CIR for breast cancer, and this trend persists across all age groups. The differences in CIR between parity categories become more pronounced with advancing age.

**Table 2.**
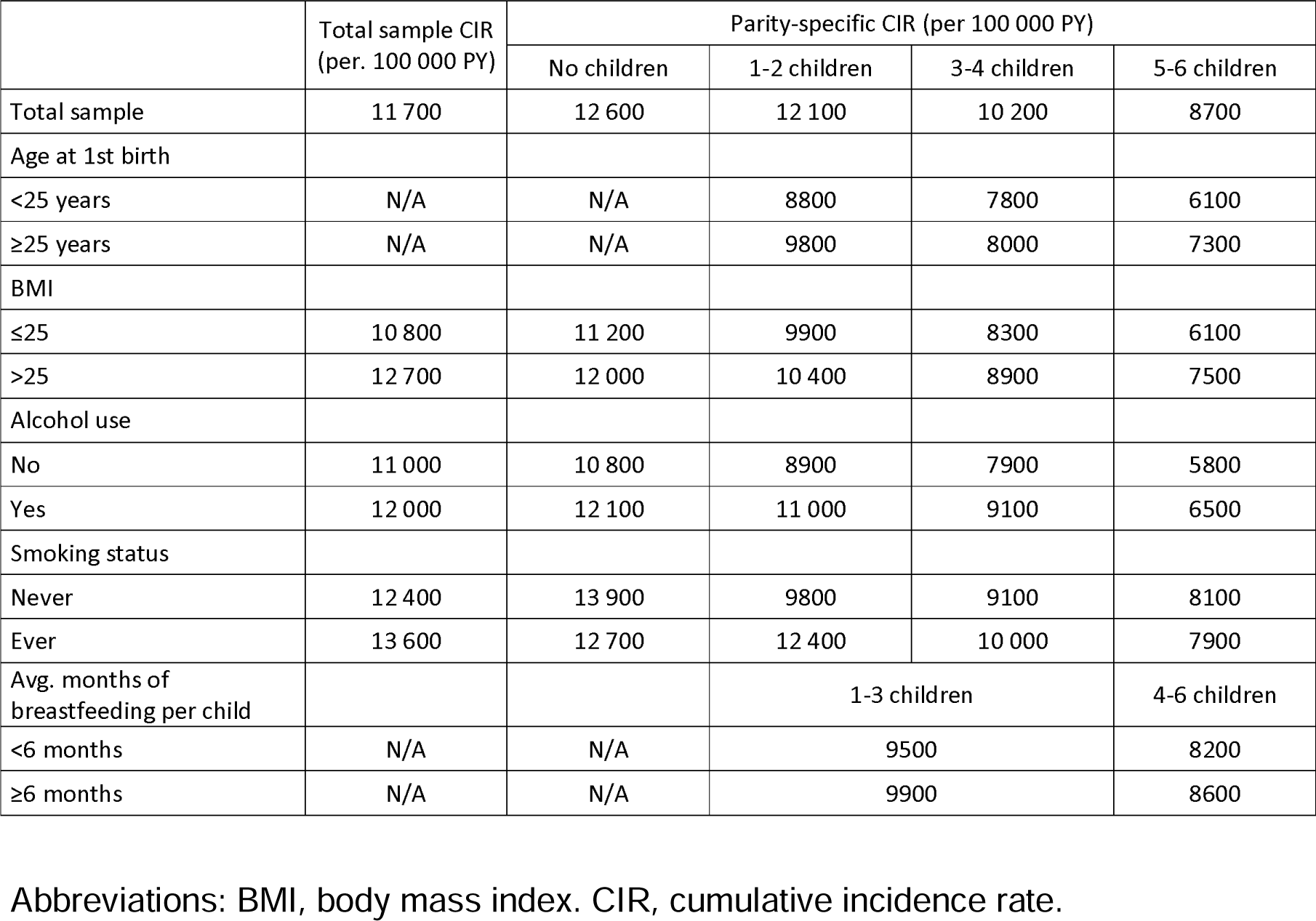
Cumulative Incidence Rates of Breast Cancer by Parity and Breast Cancer Risk Factors in the Norwegian Women and Cancer Study.

|  | Total sample CIR<br>(per. 100 000 PY) | Parity-specific CIR (per 100 000 PY) |  |  |  |
| --- | --- | --- | --- | --- | --- |
|  |  | No children | 1-2 children | 3-4 children | 5-6 children |
| Total sample | 11 700 | 12 600 | 12 100 | 10 200 | 8700 |
| Age at 1st birth |  |  |  |  |  |
| <25 years | N/A | N/A | 8800 | 7800 | 6100 |
| ≥25 years | N/A | N/A | 9800 | 8000 | 7300 |
| BMI |  |  |  |  |  |
| ≤25 | 10 800 | 11 200 | 9900 | 8300 | 6100 |
| >25 | 12 700 | 12 000 | 10 400 | 8900 | 7500 |
| Alcohol use |  |  |  |  |  |
| No | 11 000 | 10 800 | 8900 | 7900 | 5800 |
| Yes | 12 000 | 12 100 | 11 000 | 9100 | 6500 |
| Smoking status |  |  |  |  |  |
| Never | 12 400 | 13 900 | 9800 | 9100 | 8100 |
| Ever | 13 600 | 12 700 | 12 400 | 10 000 | 7900 |
| Avg. months of<br>breastfeeding per child |  |  | 1-3 children |  | 4-6 children |
| <6 months | N/A | N/A | 9500 |  | 8200 |
| ≥6 months | N/A | N/A | 9900 |  | 8600 |

Abbreviations: BMI, body mass index. CIR, cumulative incidence rate.

**Figure 2a.**
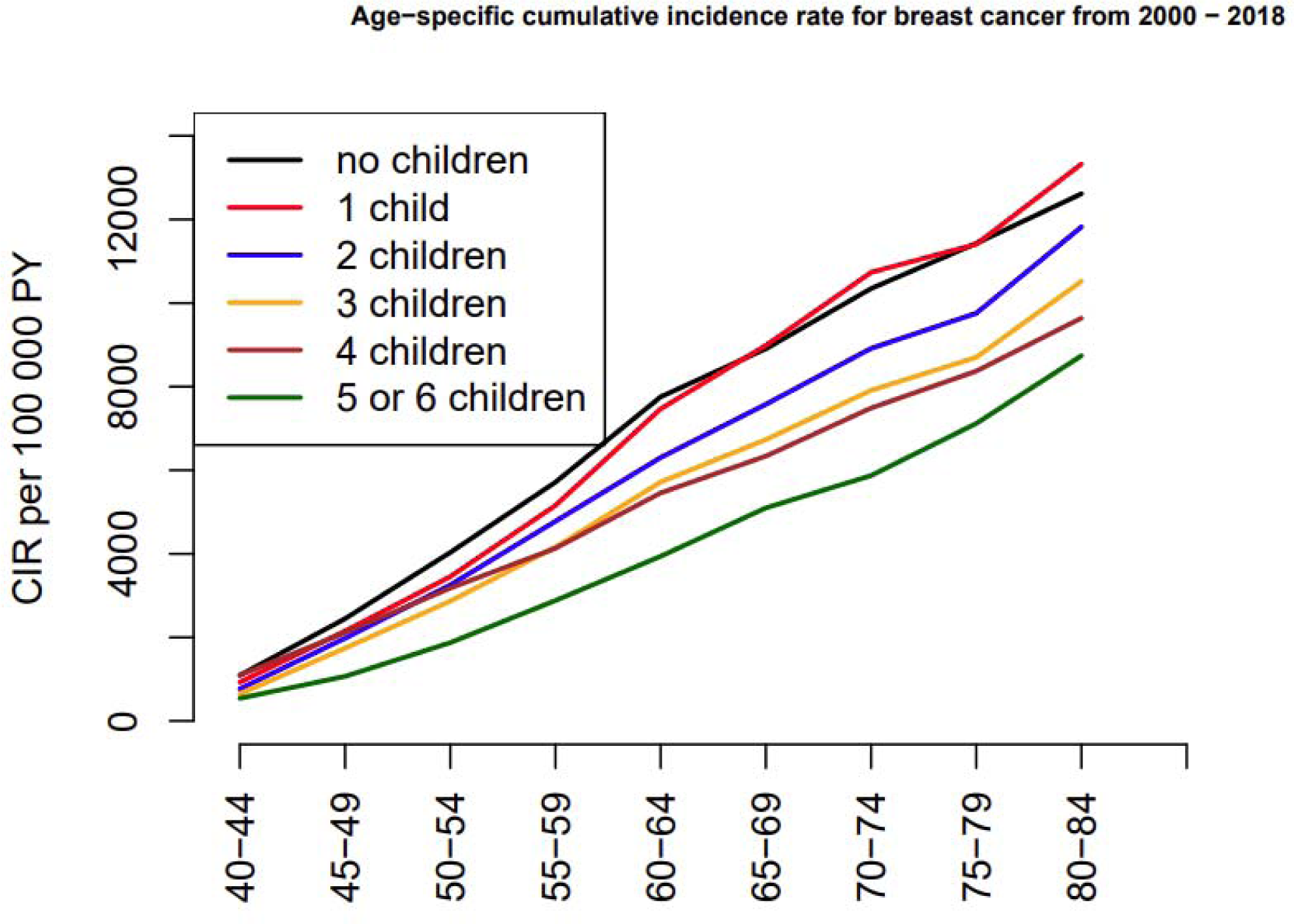
Cumulative incidence rates according to parity and parity cohorts in NOWAC 2000-2018.

**Figure 2b.**
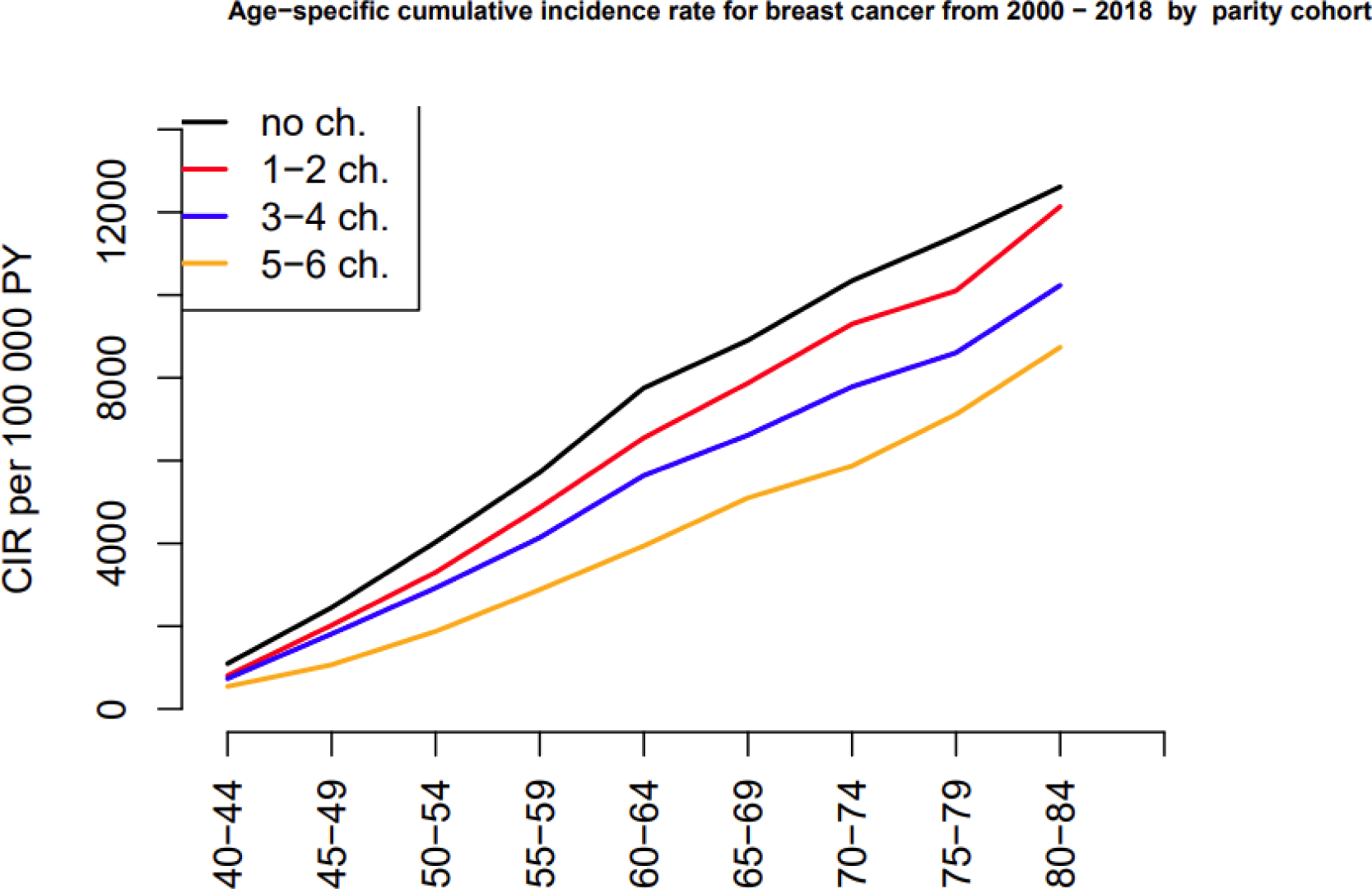
Cumulative incidence rates according to parity groups in NOWAC 2000-2018.

### 3.3 Parity-specific CIR of BC, stratified by others BC risk factors

For all analyses of parity-specific CIR of BC stratified by others BC risk factors, parity was categorized into groups. Overall, CIR of BC declines with increasing number of children, in all strata of other risk factors. Within the same parity group, women exposed to others BC risk factors had higher CIR compared to non-exposed.

#### Age at first birth

The CIR for mothers who had their first child before the age of 25 was lower in all three parity groups compared to mothers who had their first child after the age of 25 (Table 2, Figure 3). Figure 3 shows the association between parity and age-specific CIR, in both strata of age at first birth. Largely (Table 2, Figure 3), those with more children have consistently lower CIR compared to those with fewer children, regardless of age at first birth.

**Figure 3.**
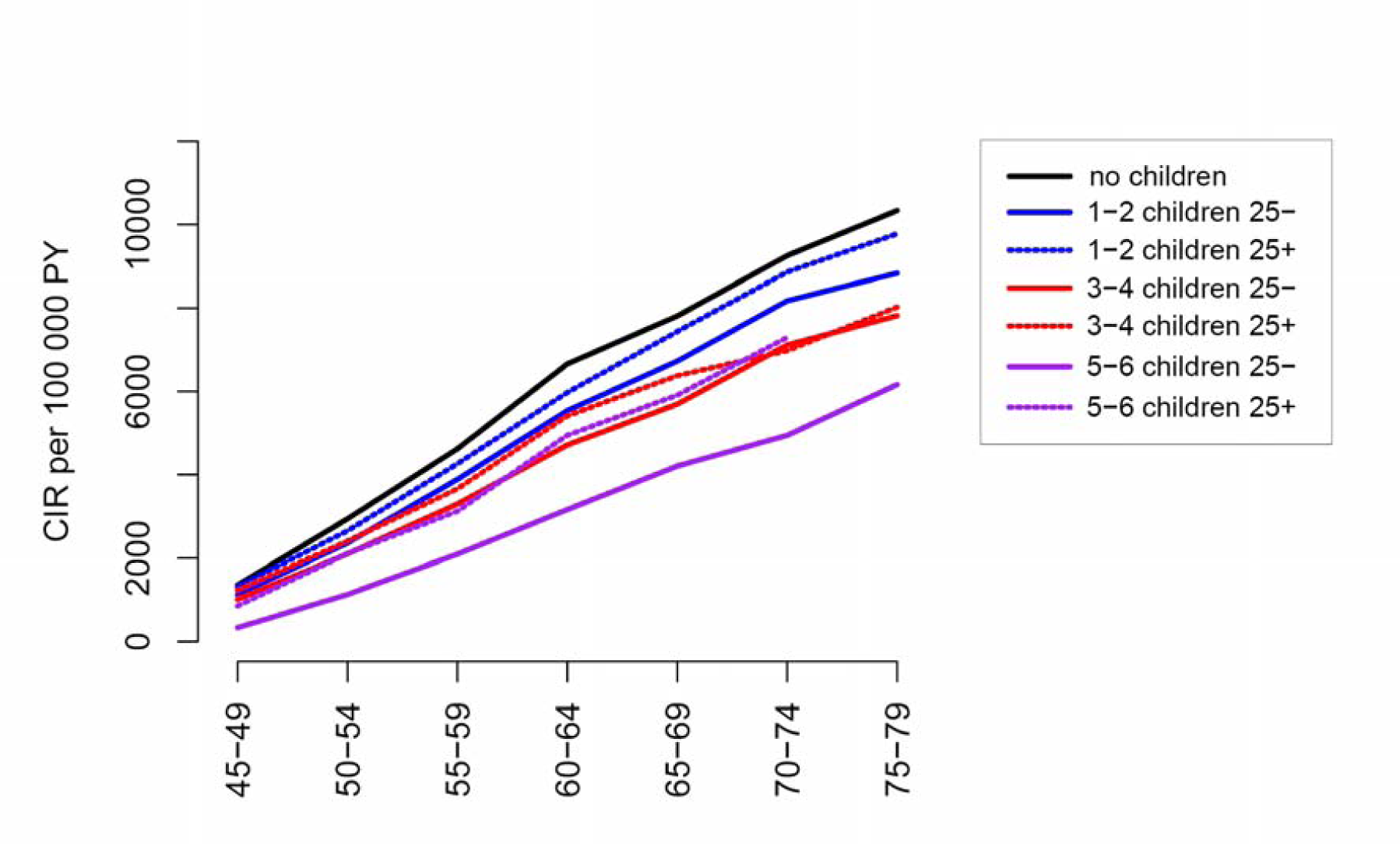
Cumulative incidence rates for breast cancer by parity, stratified by age at first birth less than 25 years, or ≥25 years.

#### Breastfeeding per child

Due to few cases in each group, analyses of breastfeeding were divided into two parity groups, 1-3 and 4-6 children, and the duration of breastfeeding was defined as below or above 6 months per child. Women with 1-3 children and longer breastfeeding duration, had a CIR of 9900 compared to 9500 per 100 000 PY for those with shorter duration (Table 2).

The pattern was the same for 4-6 children, with lower CIR in those with shorter breastfeeding duration per child (8200 versus 8600 per 100 000 PY). Figure 4 shows the association between parity and age-specific CIR, in both strata of breast feeding. In both parity groups, longer breastfeeding duration is associated with higher CIR. Overall (Table 2, Figure 4), those with 4-6 children have consistently lower CIR compared to those with 1-3 children, regardless of avg. duration of breastfeeding per child.

**Figure 4.**
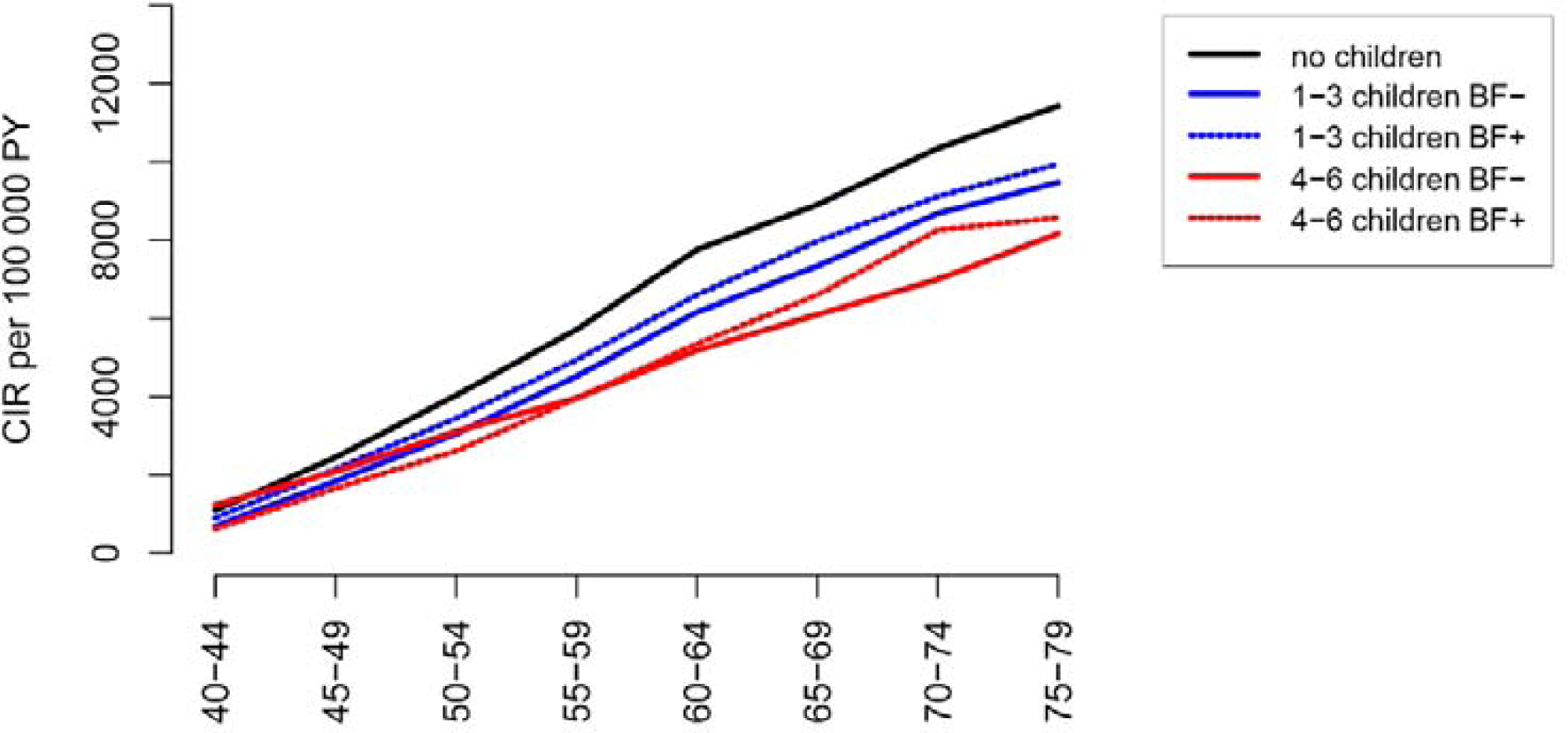
Cumulative incidence rates for breast cancer by parity, stratified by avg. duration of breast feeding for each child (more than 6 months (BF+), or shorter (BF-)).

#### BMI

In the total study sample, CIR of women with lower BMI (<25) was 10 800 compared to CIR 12 700 per 100 000 PY for women with higher BMI (≥25, Table 2, Figure 5a).

**Figure 5a.**
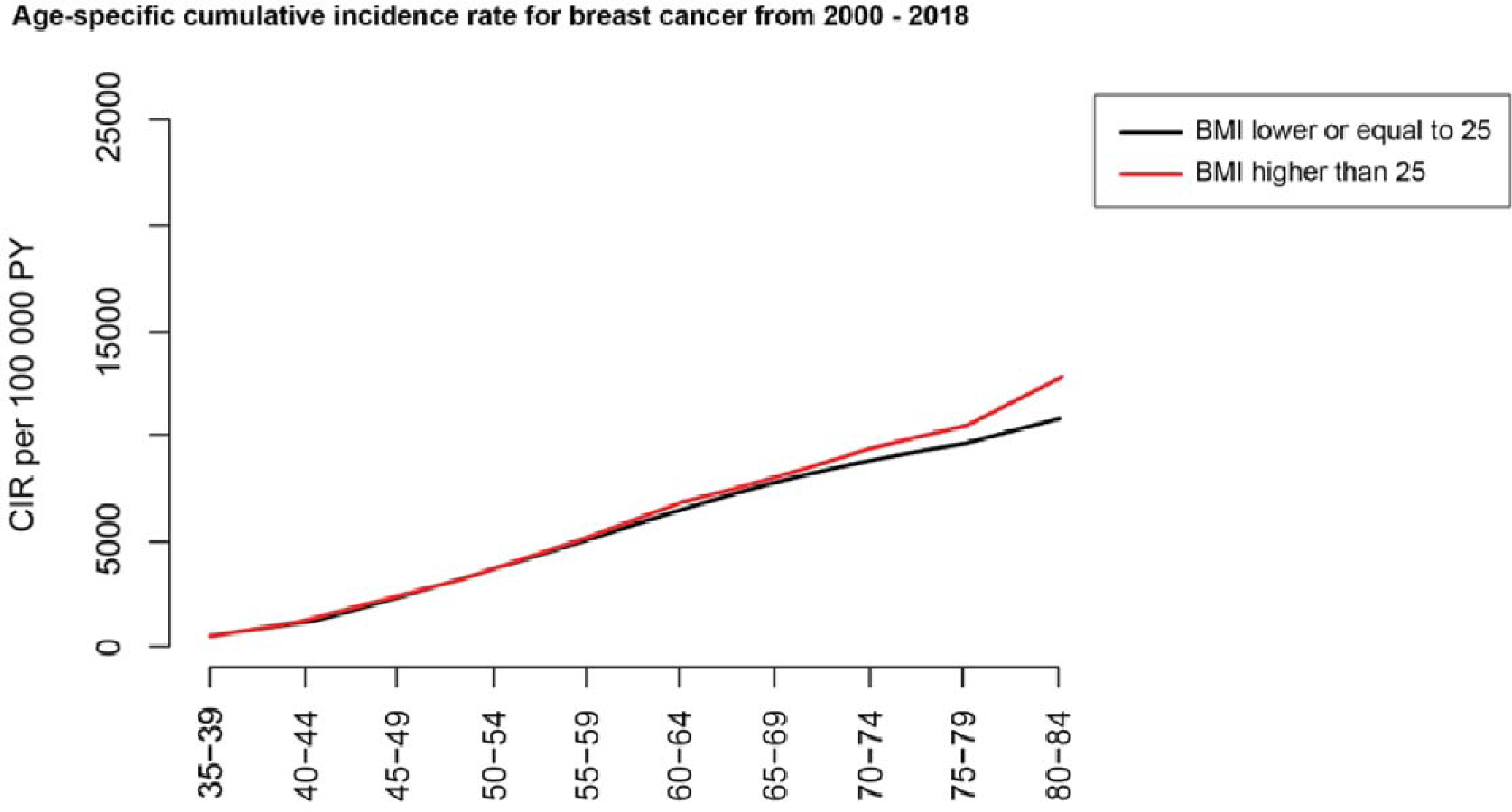
Cumulative incidence rates for breast cancer, stratified by BMI (<25 25-) or >=25 (25+).

For nulliparous women with lower BMI, CIR was 11 200 compared with CIR 12 000 per 100 000 PY for nulliparous with higher BMI (Table 2). In comparison, for those with 5-6 children and a lower BMI CIR was 6100, and 7500 per 100 000 PY for those with higher BMI. In each parity group, CIR of BC was higher for women with higher BMI.

Overall (Table 2, Figure 5b), those with more children have lower CIR compared to those with fewer children, regardless of BMI group.

**Figure 5b.**
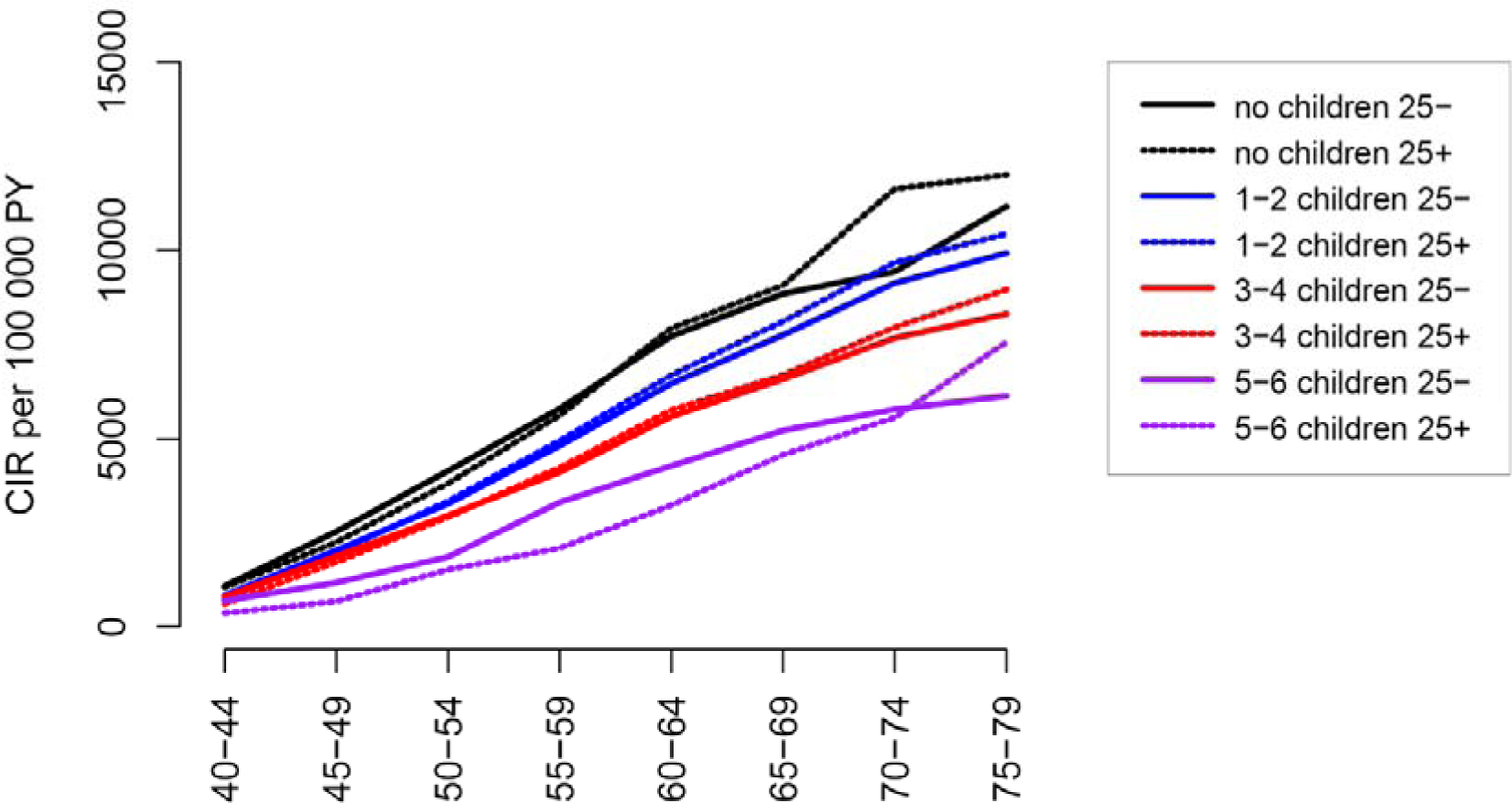
Cumulative incidence rates for breast cancer by parity, stratified by BMI (<25 (25-) or >=25 (25+).

#### Alcohol use

In the total study sample, CIR of women who did not consume alcohol was 11 000 and 12 000 per 100 000 PY for alcohol consumers (Figure 6a, Table 2).

**Figure 6a.**
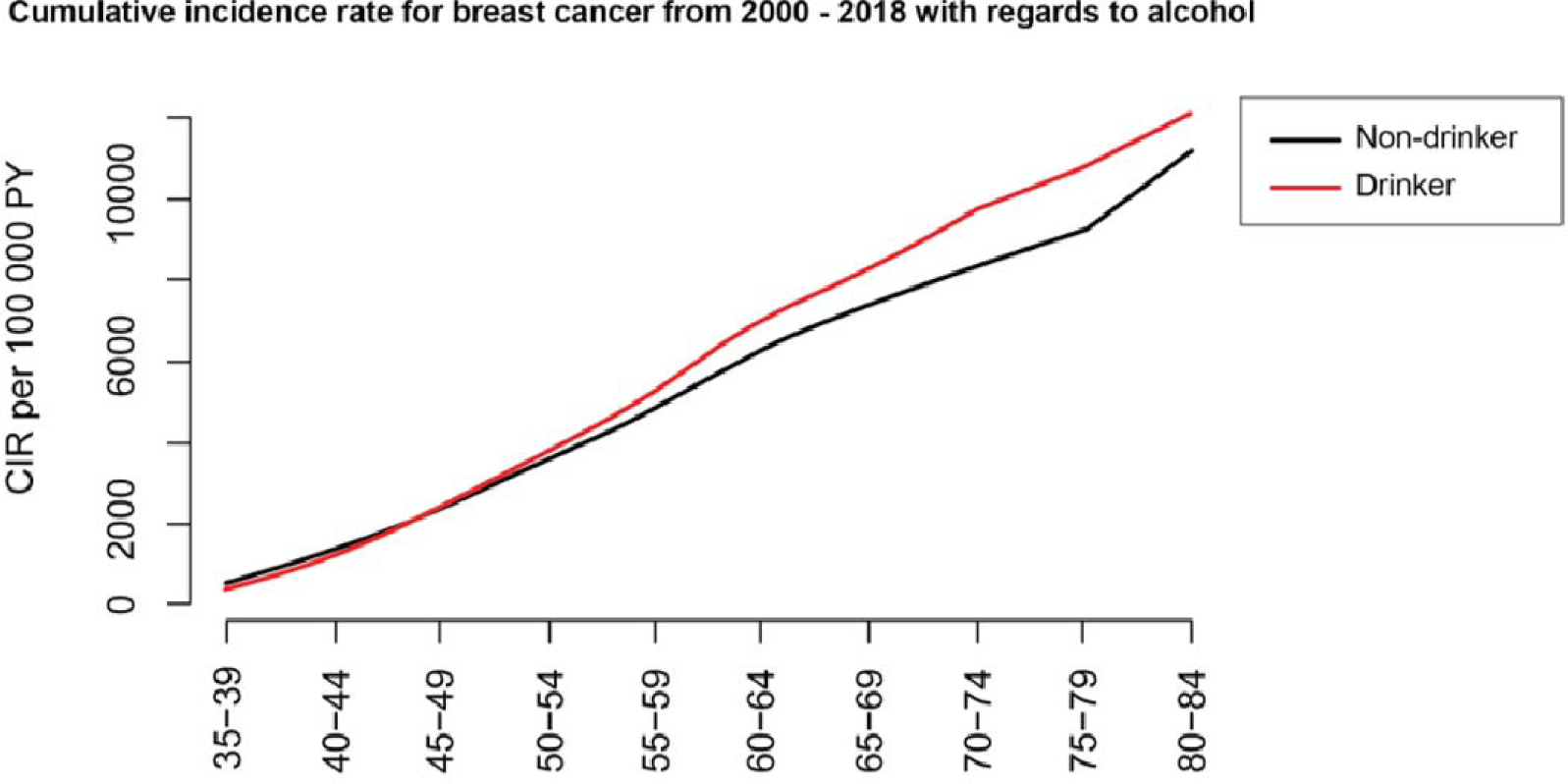
Cumulative incidence rates for breast cancer stratified by alcohol consumption (never drinking (non-d) versus alcohol consumption (drink)).

Overall, those with more children had lower CIR compared to those with fewer children, regardless of alcohol consumption status (Figure 6b, Table 2).

**Figure 6b.**
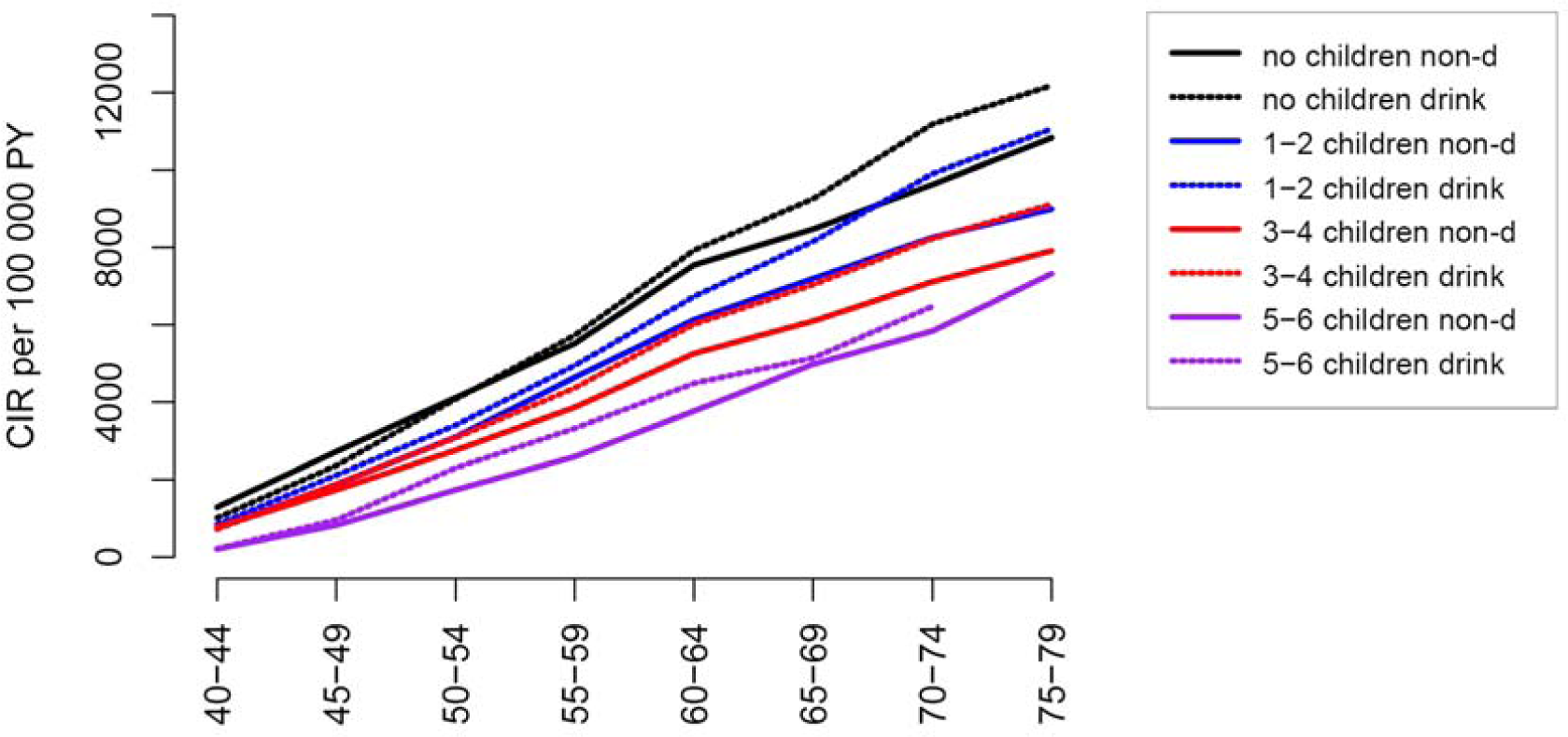
Cumulative incidence rates for breast cancer by parity, stratified by alcohol consumption (never drinking (non-d) versus alcohol consumption (drink)).

#### Smoking

CIR in the total study sample, was 12 400 for ever smokers and 13 600 per 100 000 PY for never smokers (Table 2, Figure 7a). Similarly to the analyses of the others BC risk factors, CIR was lower in those with more children compared to fewer children, regardless of smoking status (Table 2, Figure 7).

**Figure 7a.**
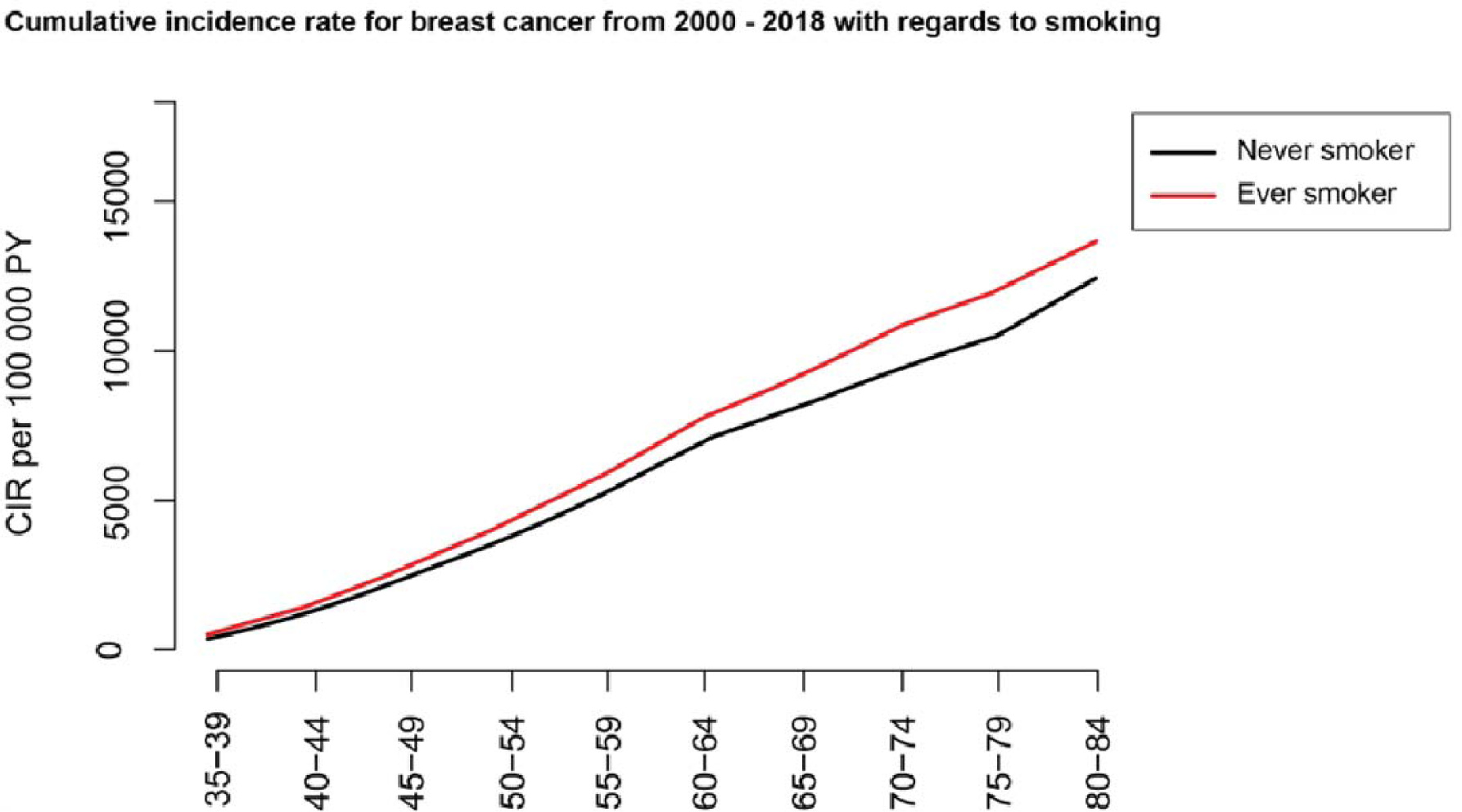
Cumulative incidence rates for breast cancer stratified by smoking (ever versus never smokers)

**Figure 7b.**
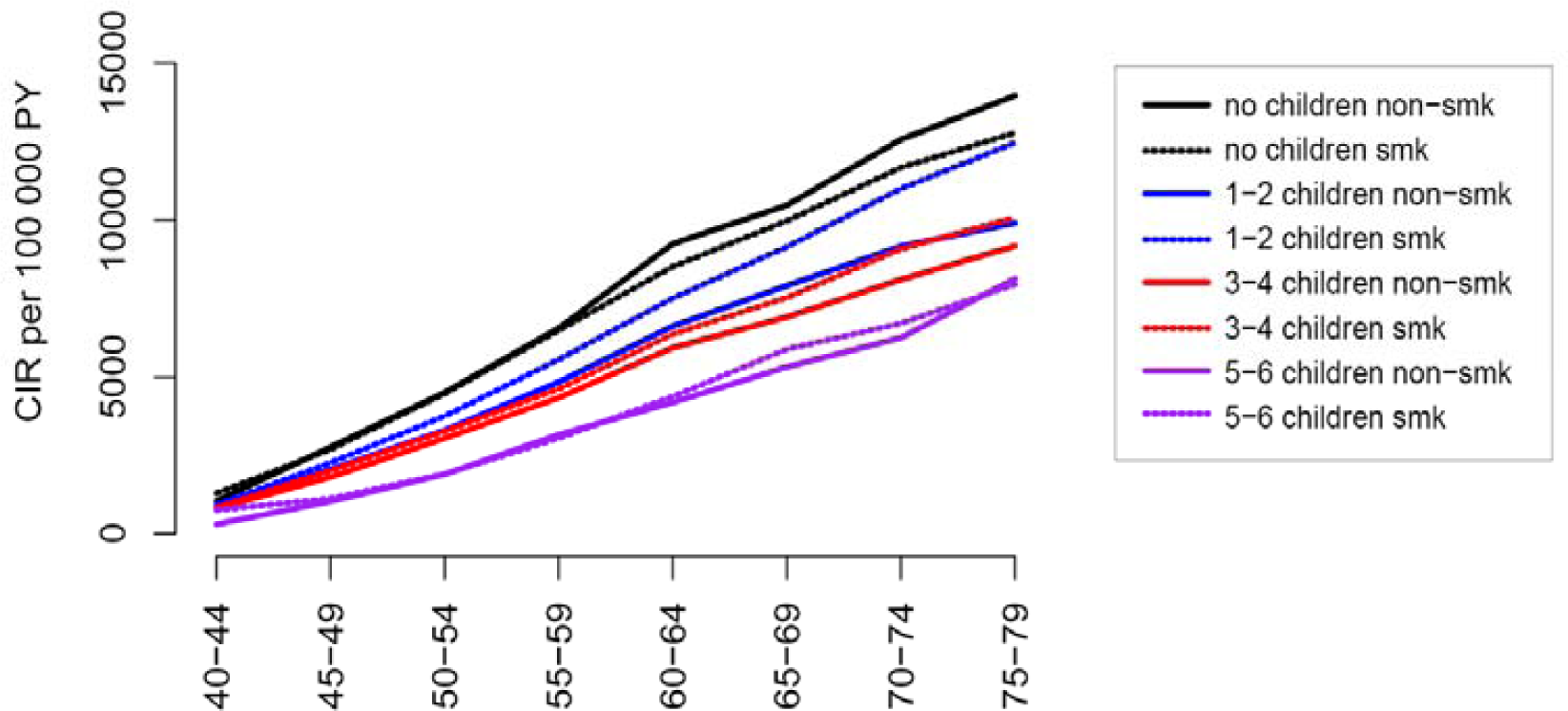
Cumulative incidence rates for breast cancer by parity, stratified by smoking (ever smokers (smk) versus never smokers (non-smk)).

## 4 Discussion

In this study, we describe the risk of BC in women 40 years and older in the form om cumulative incidence rate (CIR), and in relation to parity. Our results show an average decrease in CIR of 0.8% per child. The parity-specific CIR had the same pattern of decrease according to parity, and the risk by parity was independent of other risk factors. The associations between the parity and risk of BC found in this study are consistent with existing research.

To our knowledge, there are only a few studies using CIR as risk estimate for BC according to parity. Colditz et al., used data from the Nurses’ Health Study to calculate the cumulative risk of BC up to the age of 70 years. They showed that women with multiple births and a first birth at an early age, had a reduced risk of BC at or after menopause, relative to nulliparous women. However, they did not stratify for parity (16). A study by The Collaborative group on hormonal factors, estimated a 50% reduction of cumulative incidence of BC in developed countries, if women had the same average number of births as women in developing countries. They emphasized breastfeeding as a factor that could account for almost two-thirds of this estimated difference (13).

### 4.1 Aspects of the parity-specific CIR of BC

Reproductive history is the strongest known modifier of a woman’s breast cancer risk. This broad exposure may be comprised of a combination of being parous or not, age at first birth, number of pregnancies (full term or not), and breastfeeding (including duration), all at different levels of detail. The association between parity and risk of BC is complex. Some studies argue that BC risk is mainly determined by age at first birth and number of full-time pregnancies (9). Studies have found that multiple pregnancies reduce a woman’s risk of developing BC by 8% for each additional pregnancy (8, 9).

The potential mechanisms behind early full-term pregnancy induced protection against BC are not fully understood. Several studies, including gene expression and epigenetic studies, explored the hypothesis that first full-term pregnancy affects remodeling in mammary tissue by inducing differentiation of breast cells that make them less sensitive to carcinogenic influence (18–20) In our study, each additional childbirth reduces BC risk in a linear fashion starting from the first birth. Hence, our findings do not support the hypothesis that it is the first pregnancy that has a decisive role in the protection against postmenopausal BC.

For breast feeding duration per child, we found that longer duration was associated with higher CIR, for both women with 1-3 and 4-6 children. This finding is not in line with results from the Collaborative group {Collaborative Group on Hormonal Factors in Breast Cancer, 2002 #4582} and others. However, in our study, the number of children was a stronger predictor of CIR compared to breastfeeding duration. Breastfeeding rates in Norway were low at the end of the 1960s and the 1970. Bottle feeding was modern and promoted by formula companies. At that time, exclusive breastfeeding, beyond 4 months was not advocated (21). Hence, we postulate that the observed relationship between longer breastfeeding duration and higher CIR could be due to confounding factors. For instance, women who breastfed longer may have been a select group with different characteristics or risk factors for breast cancer. This could be part of the explanation of the different risk estimates, compared to the Collaborative group results.

Husby et al. showed that pregnancies lasting 34 weeks or longer were associated with considerable risk reduction, compared with pregnancies lasting less than 34 weeks (22). In the same study they showed that both live and stillbirths were associated with risk reduction given that the pregnancy lasted 34 weeks or longer. These overall findings strengthen the hypothesis that parity, and not breastfeeding, is the main driver of the risk reduction. The distinctive difference in risk of BC by pregnancies lasting longer or shorter than 34 weeks, suggests that biological processes that take place towards the end of the pregnancy may change the BC susceptibility of the mother. Lund et al. highlighted involvement of the immune system in parity-associated BC protection, with the use of a systems epidemiology approach (8). In sum, the protective effect of parity can be due to processes that take place locally in the breast tissue, or systemic, involving the immune system.

### 4.2 The influence of other risk factors on the parity-specific CIR of BC

Analyses for selected risk factors showed consistently higher parity-specific CIR of BC for those exposed, and exposure to risk factors did not hamper the protective effect of increasing parity. The reduction in CIR with increasing parity in most sub-strata demonstrates that the protective effect of parity is mainly independent of other risk factors.

In our total study sample, we find no difference in CIR of BC in women below 65 years of age in relation to BMI. Women aged 65 and older, with BMI ≥ 25, had a higher risk of BC compared with women in the same age group with BMI <25. Previous studies showed that higher BMI in premenopausal age is protective for BC but in postmenopausal age, higher BMI increases the risk of BC (23). Our results show that BMI had a similar influence on risk of BC in all parity groups, with the largest difference in CIR by BMI in nullipara over the age of 65, and in the highest parity group of 5-6 children. Overall, BMI had only a moderate influence on the association between parity and BC risk.

Alcohol consumption consistently increased CIR of breast cancer in the total study population, and within parity groups. This is in line with other studies that show an association between alcohol consumption and BC risk (24, 25).

Overall, smokers in our study had higher CIR than non-smokers. Stratified by parity, nulliparous women who smoked had slightly lower CIR than nullipara who did not smoke. However, we consistently find that women who have children have lower CIR than nullipara, regardless of smoking status. As for the results on breastfeeding duration, we suspect that the finding of higher risk in non-smoking nullipara women compared to smoking nullipara, may be due to confounding factors.

### 4.3 Strengths and limitation

In contrast to most of the available evidence, this study uses CIR as a measure of risk. CIR provides a more nuanced understanding of how the risk of breast cancer accumulates over a lifetime and in different age groups. Additional strengths of the current study are the prospective design and random selection of participants. To ensure a representative sample, each participant was randomly selected from the national population register, with birth year serving as the sole criterion for sample differentiation. Previous analyses have confirmed that the participants in the NOWAC study exhibit the same level of fertility as the general population (23). In addition, the external validity of the current study was demonstrated using data from the National Cancer Registry for the same period. The study is prospective with information on parity and other risk factors collected before start of follow-up. While the statistical power of the total study was high, the stratification left several analyses with few cases, especially below 40 and above 80 years of age. Hence, in the stratified analyses these age-groups were removed, and our results cannot be extrapolated to these age groups.

The external validity in terms of relevance of our findings to today’s population may be discussed. The average number of children per woman was 2.2 in the present study, whereas the Norwegian average in 2022 was 1.41 (29). Similarly, the age at first birth in this study was 23.8 years, which is considerably lower than the average of 30 years in Norway today (29). Further, the age at first birth in Norwegian women has changed substantially over the last decades. In year 2000, women aged 35-39 years gave birth to 14.2% of all children born that year. In 2018 this number had increased to 20.4.% (30). Women over 40 years of age give birth to less than 2% of children. Consequently, most of our analyses started at age 40 years to capture the effect of childbirths taking place later in life.

This study has some limitations related to the questionnaire information. Self-reported height and weight for BMI calculations may be biased. However, in a validation study in NOWAC, self-reported weight and height provided a valid ranking of BMI for middle-aged Norwegian women (31). Information on smoking and alcohol consumption was self-reported and did not include additional information on history or intensity of exposure. Smoking variables were not validated in NOWAC, but smoking status at baseline and follow-up questionnaires were compared in 2018, with only 1.8% of participants reported that they were ever smokers at baseline and never smokers at follow-up (32). A low degree of non-differential misclassification is therefore expected.

## 5 Conclusion

Our findings support the evidence that parity is a protective factor for the development of breast cancer, and highlights that this may be irrespective of other established risk factors, including those related to pregnancy (age at first birth and duration of breastfeeding) and other lifestyle factors. Notably, each childbirth, rather than only the first, is associated with a reduction in breast cancer risk among postmenopausal women.

## Data Availability

The data used in the present study may be made available from the Norwegian Women and Cancer study through their application form (www.uit.no/NOWAC).

## Declarations

### Ethical approval and consent to participate

The NOWAC study was approved by the Norwegian Data Inspectorate and the Regional Ethical Committee of North Norway/ The Regional Committee for Medical and Health research Ethics. The study was conducted in compliance with the Declaration of Helsinki, and all participants gave written informed consent. The linkages of the NOWAC database to national registries such as the Cancer Registry of Norway and registries on death and emigration were approved by the Directorate of Health. The women were informed about these linkages.

### Availability of data and material

The data may be made available from the Norwegian Women and Cancer study through their application form (www.uit.no/NOWAC).

### Authors contributions

Study design: EL. Data analysis: EL, ABW, ML and SKH. Data/results interpretation: EL, SKH, ML, KSO. Funding acquisition: EL. Writing, review and/or revision of the manuscript: SKH, EL, ABW, RHP, ML and KSO. All authors reviewed and approved the final manuscript.

## Acknowledgments

We would like to thank all participants and staff in the NOWAC study for their contributions. Some of the data in this article are from the Cancer Registry of Norway. The Cancer Registry of Norway is not responsible for the analysis or interpretation of the data presented.

## Abbreviations

NOWAC: Norwegian Women and Cancer Study
BC: Breast cancer
BMI: Body mass index
HT: hormone therapy
RR: relative risk

